# Gait and Balance Function Improves After 10 Weeks of Using a Wearable Sensory Neuroprosthesis in Persons with Peripheral Neuropathy and High Fall Risk – the walk2Wellness Trial

**DOI:** 10.1101/2020.08.02.20166231

**Authors:** Lars IE Oddsson, Teresa Bisson, Helen S Cohen, Laura Jacobs, Mohammad Khoshnoodi, Doris Kung, Lewis A Lipsitz, Brad Manor, Patricia McCracken, Yvonne Rumsey, Diane M Wrisley, Sara R Koehler-McNicholas

## Abstract

**Background:** Patients with sensory peripheral neuropathy (PN) commonly present with gait and balance problems increasing their risk of falls. The multi-site walk2Wellness trial investigates effects of long-term, home-based daily use of a wearable lower limb sensory neuroprosthesis on gait function, balance, quality of life and fall rates in a cohort of PN patients. The device (Walkasins®, RxFunction Inc., MN, USA) is designed to replace lost nerve function related to foot pressure sensation by providing directional tactile cues around the ankle reflecting foot pressure measurements during standing and walking activities. We hypothesized that previously shown short-term in-clinic improvements in gait and balance would be sustained after 10 weeks of use.

**Methods:** Participants had a PN diagnosis with loss of plantar sensation associated with gait and balance problems, a Functional Gait Assessment (FGA) score <23 (cut-off for high fall risk), and ability to sense Leg Unit tactile stimuli. Clinical outcomes included FGA, Gait Speed, Timed Up&Go (TUG) and Four-Stage Balance Test. Patient-reported outcomes included Activities-Specific Balance Confidence (ABC) scale, Vestibular Disorders Activities of Daily Living Scale (VADL), PROMIS participation and satisfaction scores, pain rating, and falls. Evaluations were performed at baseline visit and after 2, 6, and 10 weeks. Subjects were not made aware of any changes in outcomes and no additional balance interventions were allowed.

**Results:** Forty-five participants of 52 enrolled across four sites completed all in-clinic assessments. FGA scores improved from 15.0 at baseline to 19.1 at 10 weeks (p<0.000001), normal and fast gait speed from 0.86m/s to 0.95m/s (p<0.00005) and 1.24m/s to 1.33m/s (p<0.002), respectively, and TUG from 13.8s to 12.5s (p<0.012). Four-Stage Balance Test did not improve significantly. Several patient-reported outcomes were in normal range at baseline and remained largely unchanged. Interestingly, while FGA scores improved similarly across the baseline range, subjects with baseline ABC scores lower than 67% (cut-off for high fall risk) showed an increase in their ABC scores (from 49.9% to 59.3%, p<0.01), whereas subjects with baseline ABC scores above 67% showed a decrease (from 76.6% to 71.8%, p<0.019). Subjects who reported falls in the prior six months (n=25) showed a decrease in the number of fall-risk factors (from 5.1 to 4.3, p<0.023) as well as a decrease in fall rate from 13.8 to 7.4 falls/1000 days (p<0.014). Four subjects in the pre-study non-faller group (n=20) fell during the 10 weeks of the study.

**Conclusion:** A wearable sensory neuroprosthesis may provide a new way to treat gait and balance problems and manage falls in high fall-risk patients with PN.

**Trial registration:** ClinicalTrials.gov (#NCT03538756)

## 1 Introduction

Falls are a widely recognized problem in the elderly (Ganz and Latham, 2020). About 29% of community-dwelling adults 65 years or older fall once annually and 10% fall at least twice annually (Ganz et al., 2007; Bergen et al., 2016). Data from the Centers for Disease Control (CDC) indicate that medical treatment was required by 37.5% of individuals who fell in 2014 (Bergen et al., 2016). Sterling et al. (Sterling et al., 2001) reported that 30% of falls in the elderly result in serious injury. In 2015, medical cost related to fatal and nonfatal falls was approximately $50.0 billion (Florence et al., 2018). Overall, falls are associated with poor health, shortened survival (Jónsdóttir and Ruthig, 2020), reduced quality of life, and a fear of falling (Lawrence et al., 1998; Scheffer et al., 2008). Sensory peripheral neuropathy (PN) is associated with poor balance and is an independent risk factor for falls (Richardson and Hurvitz, 1995), regardless if the etiology is idiopathic (Riskowski et al., 2012), due to diabetes (Mustapa et al., 2016; Vinik et al., 2017), or chemotherapy (Winters-Stone et al., 2017). The prevalence of PN in the US population for those over age 40 has been reported to be nearly 15% (Gregg et al., 2004). The importance of sensory information from plantar cutaneous mechanoreceptors for balance control has been shown in healthy individuals (Meyer et al., 2004a; b), with loss of such information in patients with PN likely leading to problems with gait and balance function and increased risk of falls (Menz et al., 2004; DeMott et al., 2007; Dixon et al., 2017; Lipsitz et al., 2018). The occurrence of fall-related injuries is up to 15 times higher in patients with diabetic PN than in healthy individuals (Cavanagh et al., 1992). Furthermore, the prevalence of polyneuropathy has been reported to be almost four times higher in persons older than 60 years and to independently contribute to functional impairments including difficulty walking and tendency to fall (Hoffman et al., 2015). Persons with polyneuropathy are more likely to fall and more often incur fall-related injuries (Hanewinckel et al., 2017). In a prospective study, 65% of older individuals with PN fell during a one-year period and 30% reported an injury from a fall (DeMott et al., 2007). In addition, low gait speed is a risk factor for falls (Studenski et al., 2003; Montero-Odasso et al., 2005), an important indicator of frailty (Kim et al., 2019) and a predictor of survival (Studenski et al., 2011). Although gait speed declines with healthy aging (Buracchio et al., 2010), the decline in individuals with progressive sensory loss may be four times as high (Lipsitz et al., 2018). Interestingly, interventions designed to improve gait speed may also increase survival (Hardy et al., 2007).

Clinical treatment of gait and balance problems related to PN is mainly limited to the use of canes, walkers, physical therapy interventions and balance exercises (Richardson et al., 2001; Ganz and Latham, 2020) including Tai-Chi (Li and Manor, 2010; Manor et al., 2014; Quigley et al., 2014). Long-term use of bilateral ankle foot orthoses in elderly individuals with a history of falls showed positive changes in certain in-clinic static sway measures (Wang et al., 2019a), although long-term benefits related to fall rates and gait function appear limited (Wang et al., 2019b).

Several review studies support the hypothesis that strength and balance training interventions can improve balance and reduce fall risk and falls in patients with PN (Ites et al., 2011; Tofthagen et al., 2012; Streckmann et al., 2014). The training, however, should be specific to balance (Bulat et al., 2007; Oddsson et al., 2007; Halvarsson et al., 2011; Akbari et al., 2012) because strength and/or endurance training in patients with PN appears to have less impact on balance (Streckmann et al., 2014). In addition, unless balance activities, including Tai Chi or balance therapies are conducted with sufficient intensity, frequency (Lipsitz et al., 2019) and specificity, benefits may be limited or absent (Kruse et al., 2010; Lipsitz et al., 2018; Lipsitz et al., 2019) leading to mixed outcomes. Furthermore, continued exercise is required to maintain benefits long-term (Wolf et al., 2001; Halvarsson et al., 2013; Melzer and Oddsson, 2013), although some improvements last up to six months (Allet et al., 2010). Guidelines regarding physical activity for older adults with mobility problems recommend a minimum of activity at least twice a week (Chodzko-Zajko et al., 2009). Some studies on patients with diabetic PN following a period of balance training 2-3 times /week over 6-12 weeks did show improved balance and reduced fall risk (Morrison et al., 2010; Morrison et al., 2018). However, there currently are no specific guidelines regarding frequency of balance exercises and even three times a week may be insufficient to see an improvement in balance function (Kruse et al., 2010). Consequently, there is a need for additional solutions to help improve gait and balance function in patients with PN.

A growing body of literature on various sensory substitution and augmentation technologies suggest novel ways of improving gait and balance function in different populations of patients. The concept of sensory substitution related to brain plasticity was laid out by Bach-y-Rita and colleagues, initially for vision and the vestibular system (Bach-y-Rita et al., 1969; Bach-y-Rita, 2004) and other sensory systems (Tyler et al., 2003; Bach-y-Rita, 2004). Recent efforts include wearable systems showing benefits to patients with vestibular loss (Hegeman et al., 2005; Wall et al., 2009; Basta et al., 2011; Yamanaka et al., 2016), PN (Wall et al., 2012; Wrisley et al., 2018) and Parkinson’s Disease (Rossi- Izquierdo et al., 2013; Lee et al., 2015). Combining wearable neurostimulation with balance therapy has shown benefits in patients with multiple sclerosis (Leonard et al., 2017), cerebellar ataxia (Cakrt et al., 2012), stroke (Badke et al., 2011) traumatic brain injury (Ptito et al., 2020) and in-home balance therapy (Bao et al., 2018)

In a randomized crossover trial, a recent study further supported findings from an earlier pilot study (Wall et al., 2012) and demonstrated meaningful short-term, in-clinic improvements in Functional Gait Assessment (FGA) scores and gait speed in subjects with PN using a wearable sensory neuroprosthesis (Koehler-McNicholas et al., 2019). The device (Walkasins®, RxFunction Inc., MN, USA, Figure 1) is an external lower limb sensory prosthesis designed to replace lost nerve function used for detection and signaling of foot pressure sensation in patients with PN. It provides gentle directional tactile stimuli (in the form of low-intensity vibrations) around the lower leg that reflect changes in foot pressure distribution measured with an instrumented Foot Pad in the shoe. The subject’s nervous system senses these new tactile cues and incorporates them to improve gait and balance. Currently, effects of long-term daily use of Walkasins on clinical outcomes are unknown. The multi-site clinical trial, walk2Wellness, (NCT #03538756, www.clinicaltrials.gov) investigates long-term, home-based use of Walkasins on clinical and patient-reported outcomes of balance and gait function, quality of life, physical activity, social participation, pain and fall rates. Data from the primary endpoint of the study at 10 weeks are reported here. We hypothesized that previously demonstrated short-term in-clinic improvements in FGA score by at least four points would be sustained long-term (Beninato et al., 2014; Koehler-McNicholas et al., 2019). Early data from the trial were presented in abstract form (Oddsson et al., 2019; Oddsson et al., 2020a).

**Figure 1.**
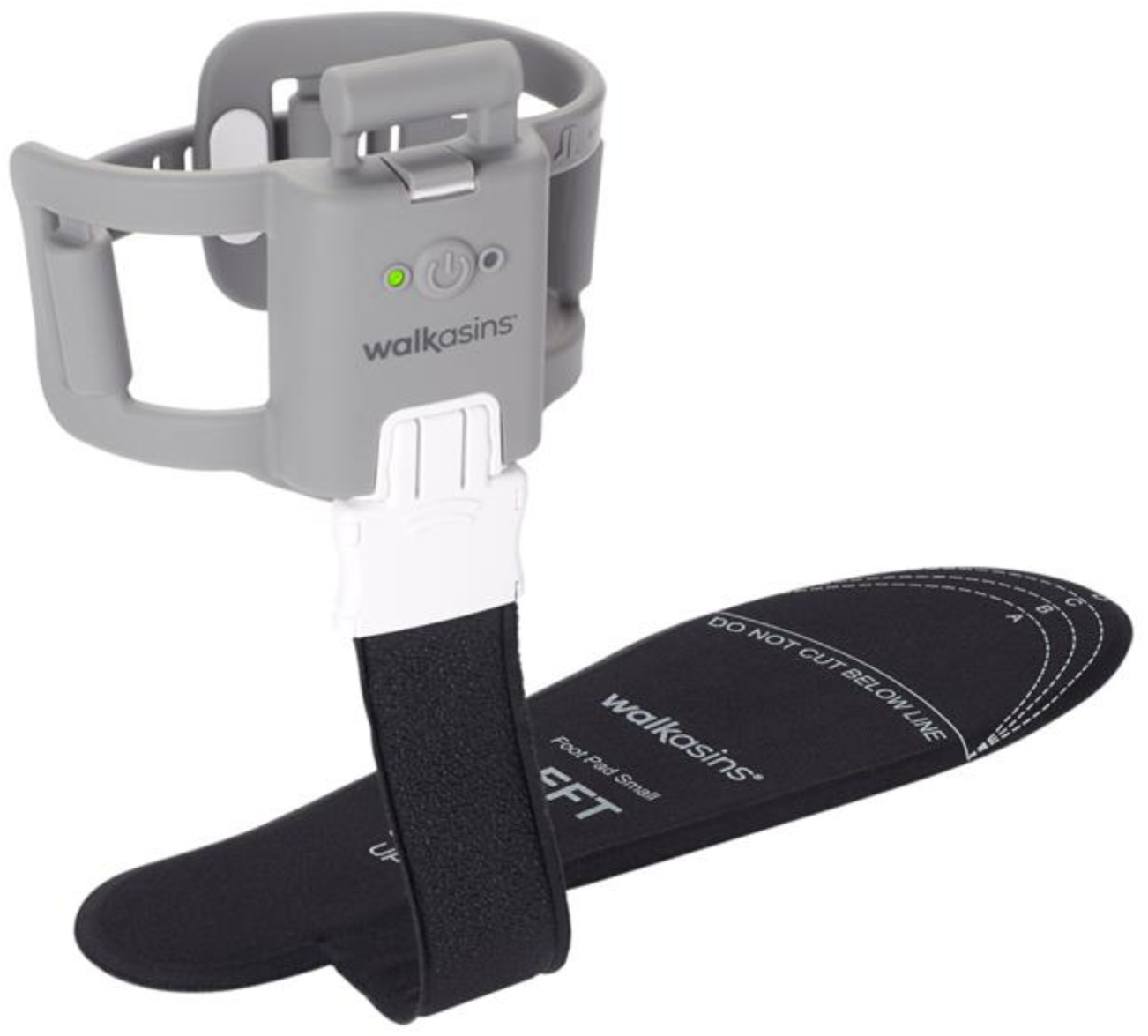
Picture of the Walkasins sensory neuroprosthesis device showing the pressure sensor embedded Foot Pad that is placed in the subject’s shoe and connected to the Leg Unit that houses an embedded microprocessor with software, supporting electronics, a rechargeable battery, and four tactile stimulators placed around the lower leg. The system is worn bilaterally. Leg Unit and left Foot Pad shown

## 2 Materials and Methods

### 2.1 Subject Recruitment

Human subject testing was approved according to the Declaration of Helsinki by Advarra IRB (formerly Quorum Review IRB), serving as the Institutional Review Board (IRB) of record for three of the participating sites under the study protocol for *walk2Wellness: Long-term Use Effects of Walkasins® Wearable Sensory Prosthesis on Gait Function, Balance-Confidence, and Social Participation*. The three sites include Baylor College of Medicine, Houston, TX; Hebrew SeniorLife, a Harvard Medical School Affiliate, Boston, MA; and M Health Fairview, Minneapolis, MN. Advarra IRB determined that Walkasins are a non-significant risk device because they do not meet the criteria of a significant risk device according to U.S. Food and Drug Administration regulations. The IRB Subcommittee, the Subcommittee on Research Safety, and the Research and Development Committee of the Minneapolis VA Health Care System (MVAHCS) also approved the trial. The study is registered on ClinicalTrials.gov (#NCT03538756). At the time this study began, Walkasins were available only for research purposes.

### 2.2 Inclusion and Exclusion Criteria

Inclusion criteria for the study were similar to Koehler-McNicholas et al. (Koehler-McNicholas et al., 2019): age 21-90 years; male or female; a formal diagnosis of sensory PN prior to participating in the study as indicated by subject’s medical record or a signed letter by a physician; self-reported problems with balance; ability for transfers or ambulation on level surfaces at fixed cadence as assessed by trained study personnel; an Functional Gait Assessment (FGA) score <23, the cut-off score for high fall risk (Wrisley and Kumar, 2010); ability to understand and provide informed consent; foot size to allow the Walkasins device to function properly, and ability to complete all functional outcome measures without the use of an assistive device to ensure sufficient motor function. Subjects could use an assistive device at their own discretion during the trial. Subjects were excluded from participation if they were unable to perceive tactile stimuli from the Walkasins leg unit or used an ankle-foot orthosis for ambulation that prevented donning of the device. Subjects with any of the following conditions were also excluded: acute thrombophlebitis; deep vein thrombosis; untreated lymphedema; a lesion of any kind, swelling, infection, inflamed area of skin, or eruptions on the lower leg near placement of the device; foot or ankle fractures; or severe peripheral vascular disease. In addition, subjects with any musculoskeletal or other neurological conditions that would prohibit use of the device, as determined by a clinician, were excluded. Due to risk of overloading the pressure sensor Foot Pad, subjects weighing over 136 kg (300lbs) were excluded from participation. Furthermore, subjects were prohibited from initiating any balance training (e.g., Tai-Chi etc.) or balance-related therapy during the ten weeks of the trial. Subjects were not systematically provided information about changes in any outcomes scores or changes in their performance throughout the 10 weeks of the trial, nor did study personnel monitor outcomes during the study. Potential subjects responded to announcements that specifically targeted individuals with PN and balance problems, or they were referred by clinicians who were familiar with the study and believed them to be good candidates for the trial.

### 2.3 Study Procedures

All participants signed IRB-approved consent forms prior to the initiation of study activities. Following the informed consent process, a study team member tested the subjects on both legs to determine whether they could feel the stimuli from the Walkasins Leg Unit (Figure 1). Subjects wore Walkasins on both feet. A small number of subjects who were unable to perceive the stimuli from the Walkasins Leg Unit (Figure 1) were excluded from participation in the study. Participants then completed a demographics and health screening questionnaire to assess common health issues related to neurological, musculoskeletal, cardiopulmonary disorders, and other systemic diseases along with information on their history of falls over the past six and twelve months and regular use of an assistive device (Koehler-McNicholas et al., 2019). Falls were defined according to the World Health Organization: “an event which results in a person coming to rest inadvertently on the ground or floor or other lower level”. Subjects enrolled in the study were instructed to wear the device as much as possible throughout their daily activities, indoors as well as outdoors. At each follow-up visit, participants were asked about changes in their health status and any falls and adverse events they experienced since the previous visit. During the baseline visit, participants also provided a list of their medications (medication name, indication, dose, and frequency), which was updated over the course of the study. Medications are a known risk factor for falling, based on side effects of medication use or drug interactions (Woolcott et al., 2009).

### 2.4 Initial Assessments

Subjects then completed the Activities-Specific Balance Confidence (ABC) Questionnaire, which measures levels of balance confidence in elderly persons. The ABC asks the question “How confident are you that you will not lose your balance or become unsteady” when performing 16 different tasks (Powell and Myers, 1995). Subjects rated themselves on a scale from 0 to 100, and an average score was calculated across the 16 responses. An ABC score below 67% has been associated with high fall risk (Lajoie and Gallagher, 2004). In addition, subjects completed the Vestibular Activities of Daily Living Scale (VADL) (Cohen et al., 2000), which evaluates self-reported effects of vertigo and balance disorders on independence in everyday activities of daily living that are relevant for individuals living in the community. Individuals rate their level of functional ability for basic and instrumental activities of daily living on a scale from 1 (independent) to 10 (dependent), which incorporates the use of assistive devices.

Following completion of the questionnaires, a study team member performed tactile and vibration sensation testing to document loss of sensation. Loss of sensation was tested with the Weinstein Enhanced Sensory Test (WEST) monofilament foot test (0.5g, 2g, 10g, 50g, and 200g) applied perpendicular to the skin at four test sites on the plantar surface of the foot, including the first, third, and fifth metatarsal heads as well as the great toe. Study personnel began testing with the 10g filament and used a smaller filament if the subject was sensate and a larger filament if the subject was insensate. The smallest filament the subject was able to feel was recorded (if none were felt this result was recorded as “none”). Vibration sensation was assessed with a Rydel-Seiffer tuning fork, which is a 128Hz tuning fork with end weights that convert the tuning fork from 128 to 64 Hz. The weights are scaled allowing a score 0-8 (lower scores indicating less sensation), allowing reliable quantitative vibratory testing. Scores were read from the black triangle and rounded to the nearest whole number. Vibration values ≤ 4 are categorized as abnormal at the first metatarsal joint (Kästenbauer et al., 2004). The tuning fork was applied firmly and perpendicular to the lateral aspect of the first metatarsophalangeal, lateral malleolus, and patella testing sites. The monofilament and vibration tests were repeated at the 10-week visit.

### 2.5 Clinical Outcome Measures

Upon completion of the monofilament and vibration sensation testing, subjects performed a series of functional outcome measures while wearing the device turned off (baseline). Tests were repeated at weeks 2, 6, and 10. Subjects could rest as needed during the clinical assessments. For study purposes the clinical outcomes were standardized and performed by study personnel who were trained by one of the investigators (DW). Observation visits were conducted periodically during the study to ensure standardization among the sites.

#### Functional Gait Assessment (FGA)

The FGA (Wrisley et al., 2004) is the recommended clinical outcome measure for walking balance based on current physical therapy Clinical Practice Guidelines for outcome measures for adults with neurologic conditions (Moore et al., 2018). It is a reliable and valid measure of gait function related to postural stability and has been shown to be effective in classifying fall risk in older adults and predicting unexplained falls in community-dwelling older adults (scores ≤22/30) (Wrisley et al., 2004; Wrisley and Kumar, 2010). It has also been validated in multiple neurological conditions (stroke, Parkinson’s, vestibular conditions) (Lin et al., 2010; Leddy et al., 2011) and has less floor and ceiling effects than the similar Dynamic Gait Index (Lin et al., 2010). The FGA includes 10 different items that challenge gait balance where each item is scored from 0 to 3 (3 = normal, 2 = mild impairment, 1 = moderate impairment, 0 = severe impairment) with a maximum score of 30. An increase of ≥4 points is considered the Minimal Clinically Important Difference (MCID) for community-dwelling elderly individuals (Beninato et al., 2014). Subjects whose baseline FGA score was 23 or higher were excluded from further participation in the study. Subjects completed the FGA in a large open area with a 6-m (20-ft) walkway marked with tape according to published recommendations (Wrisley et al., 2004).

#### 10-Meter Walk Test (10MWT)

The 10m-walk (Perera et al., 2006) is the recommended clinical outcome measure for walking speed based on current physical therapy Clinical Practice Guidelines for outcome measures for adults with neurologic conditions (Moore et al., 2018). It is routinely used in rehabilitation and has excellent reliability in multiple neurologic conditions (chronic stroke, traumatic brain injury, Parkinson’s) (Hiengkaew et al., 2012). Gait speed has been found to be an important predictor of survival in older adults (Hardy et al., 2007), further emphasizing its importance as a clinical outcomes measure. Gait speed was assessed during the middle 6 meters of a 10-meter-long pathway to allow for acceleration and deceleration, completed in one trial under two conditions: 1) walk at normal speed and 2) walk as fast as possible. An increase by 0.05 m/s is deemed “small meaningful” and 0.10 m/s as “substantial” (Perera et al., 2006). These are considered the MCID in the geriatric population (Perera et al., 2006).

#### Timed Up and Go (TUG)

The TUG (Mathias et al., 1986) is part of the CDC recommended STEADI test protocol for balance function (CDC, 2017). It is commonly used in rehabilitation and has excellent validity and reliability for elderly adults and has been shown to be effective in classifying community dwelling adults at risk for falls (Podsiadlo and Richardson, 1991; Shumway-Cook et al., 2000; Bischoff et al., 2003; CDC, 2017). From a seated position in a standard armchair, the subject is asked to do the following: 1) stand up from the chair, 2) walk at normal pace around a tape mark on the floor 10 feet from the chair, 3) turn, 4) walk back to the chair at a normal pace, and 5) sit down again. Subjects were provided one practice trial that was not recorded followed by the recorded timed trial. The tester recorded the time from the command “Go” until the subject’s buttocks returned to the chair when sitting. We used >12s as a cut-off for high fall risk (Bischoff et al., 2003; CDC, 2017). The Minimal Detectable Change (MDC) for older adults with type 2 diabetes has been reported to be 1s (Alfonso-Rosa et al., 2014).

#### 4-Stage Balance Test

The 4-Stage Balance Test is part of the Centers for Disease Control and Prevention (CDC)-recommended STEADI test protocol for balance function (CDC, 2017). It includes four gradually more challenging postures the subject is exposed to: 1) stand with feet side by side, 2) stand with feet in semi-tandem stance, 3) stand with feet in tandem stance, and 4) stand on one leg. Subjects were allowed upper extremity support to obtain the position and passed each level if they were able to hold the stance unsupported for 10 seconds. The assessment ended when subjects were unable to hold a stance for 10 seconds. The times for each position held was recorded and summed as a measure of overall performance. A fail of stances 1, 2, or 3 (i.e., total time < 30s) indicates high risk of falling (CDC, 2017).

### 2.6 Learning Protocol

As part of the baseline visit, subjects performed a standardized set of balance activities, once while wearing the device turned off and once while wearing it turned on (Koehler-McNicholas et al., 2019). Activities lasted approximately 10 minutes and included standing (two-leg standing, tandem standing, and one-legged standing) and walking (walking straight, turning right and left) at both normal and fast speeds. Activities were repeated with the eyes closed. During standing exercises subjects were challenged to explore their base of support in both mediolateral and anteroposterior directions and to notice the pattern of tactile stimuli when the device was turned on. During walking activities subjects were instructed to notice the pattern of tactile stimuli when the foot was in contact with the ground, how it matched their pace of walking, and the flow of tactile stimuli from step to step. Subjects were not instructed how to respond to the tactile stimuli from Walkasins instead, the activities focused on orientation and familiarization with the device.

### 2.7 Participant Reported Outcomes

At the baseline visit and at each follow-up visit at weeks 2, 6, and 10, subjects also completed the five subject-reported outcome measures described below:

#### Patient Health Questionnaire (PHQ-9)

The PHQ-9 (Kroenke et al., 2001) is a concise, self-administered tool for assessing depression. Commonly used for screening and diagnosis of depression, the PHQ-9 incorporates depression criteria according to the 4^th^ edition of the Diagnostic and Statistical Manual of Mental Disorders (DSM-IV) with other leading major depressive symptoms.

#### PROMIS Pain Interference Short Form 6b

(Askew et al., 2016): The PROMIS Pain Interference instrument measures the self-reported impact of pain on relevant aspects of a person’s life within the past seven days. Items capture the extent to which pain hinders social, cognitive, emotional, physical, and recreational activities. The Pain Interference short form is a global scale rather than disease specific.

#### PROMIS Numeric Rating Scale v1.0 - Pain Intensity Form 1a

The PROMIS Pain Intensity instrument assesses reported average pain intensity on a scale from 0-10 with higher scores indicating greater levels of pains. The Pain Intensity short form is global rather than disease specific.

#### PROMIS Ability to Participate Short Form 8a

The PROMIS Ability to Participate in Social Roles and Activities instrument (Hahn et al., 2016b) assesses the individual’s perceived ability to perform usual social roles and activities. The measure does not use a designated time frame (e.g. over the past seven days), and higher scores represent fewer limitations (e.g., I have trouble doing all of my regular leisure activities with others).

#### PROMIS Satisfaction with Participation in Social Roles Short Form 8a

The PROMIS Satisfaction with Social Roles and Activities (Hahn et al., 2014; Hahn et al., 2016a) is a self-reported instrument to assess satisfaction with the ability to perform usual social roles and activities (e.g., “I am satisfied with my ability to do things for my family”). PROMIS scores are presented as T-scores, a standardized score with a mean of 50 (representing average for the US population) and a standard deviation of 10.

#### User Experience Survey

At the 2 and 10-week visits, subjects completed a 10-question survey to collect information concerning their experience with the device. Subjects rated aspects of Walkasins use (e.g., donning and doffing, charging, etc.) on a 7-point Likert scale ranging from “Very Easy” to “Very Hard”. Subjects also rated their overall satisfaction with the device and were able to provide additional comments and feedback regarding their experience with the device. Between visits, subjects were asked to document their use of Walkasins on a calendar by marking the days they wore their Walkasins and for how many hours. The subject calendar was also used to facilitate documentation of falls that occurred over the course of the study (Hannan et al., 2010). Subjects were asked to return their calendars at their next study visit.

### 2.8 Number of Subjects

Sample size estimation was based on data from the recent study of a similar population of subjects (Koehler-McNicholas et al., 2019), showing a baseline average FGA score of 15.2 and standard deviation 4.8. The data was normally distributed according to the Shapiro-Wilk’s test. To detect a mean difference in pre- and post-FGA score ≥4 points, the Minimal Clinically Important Difference for community dwelling elderly individuals (Beninato et al., 2014), required at least 20 subjects using a significance level of 0.01 and a power of 0.8. Accounting for an expected ∼20% drop-out rate (National Heart, 2020) target enrollment was set at 25 subjects (20/0.8=25). Multiple sites were engaged in the trial to expand geographical, ethnical, and clinical variation in the data initially allowing each site to recruit up to 25 subjects. Due to the COVID-19 pandemic, the trial was interrupted and continued enrollment as well as in-clinic testing was halted. At this time, sufficient overall statistical power based on the sample size calculation above has been achieved following enrollment of 52 subjects, well over the 20 subjects required to achieve statistical significance. Collection of participant-reported outcomes has continued through phone calls and the longer-term outcomes assessments as originally planned (at 26 and 52 weeks) are expected to continue.

### 2.9 Statistical Analysis and Availability of Data

Descriptive statistics were calculated and presented as mean and standard deviation of the mean. Variables were tested for normality using the Shapiro-Wilk’s test. The two-proportion Z-test was used to compare proportion-based measures. Subjects who reported falls in the previous six months (Pre-Fallers, n=30) were analyzed separately from the remaining subjects (Pre-NonFallers, n=22). Comparisons of baseline characteristics between Pre-Fallers and Pre-NonFallers were made with a t-test for independent samples or a Mann-Whitney U test if data was not normally distributed based on a Shapiro-Wilk’s test. Repeated measures analysis of variance (ANOVA) was performed for outcomes measured across the four assessment events, baseline, 2, 6 and 10 weeks. If the ANOVA was significant (p<0.05), three pairwise comparisons were made using dependent t-tests between the baseline assessments and each of the 2, 6 and 10-week assessments. If the ANOVA was non-significant, no further comparisons were made. A Bonferroni’s adjustment of significance levels for correlated measures was applied, ranging from p<0.0167 (0.05/3 for three comparisons) for a full correction (non-correlated measures, r=0) and p<0.05 for perfectly correlated measures (r=1) (Uitenbroek, 1997). Effect sizes were calculated using Cohen’s d (Lakens, 2013) and were interpreted according to recommendations by Cohen (Cohen, 1988) with 0.2 representing a small effect, 0.5 a medium effect, and 0.8 a large effect. Ninety-five percent confidence intervals of effect sizes were estimated according to Algina (Algina et al., 2005). Statistical analysis was performed using the Analysis-ToolPak module in Microsoft Excel 2016 and the Real Statistics Resource Pack software, release 6.8 (Zaiontz, 2020). Table 1 shows baseline characteristics of subjects enrolled in the study from the four different clinical sites. Subject data were pooled for the continued analysis presented here.

**Table 1.** Baseline characteristics of subjects from the four different clinical sites enrolled in the study. Values represent Mean (Standard Deviation). #ChrD - Number of Chronic Diseases.

|  | n | Age<br>(yrs) | Height<br>(m) | Weight<br>(kg) | #ChrD | FGA<br>Score | Gait Speed<br>Normal<br>(m/s) | Gait Speed<br>Fast<br>(m/s) | TUG<br>(s) | 4-Stage<br>Balance<br>Test (s) |
| --- | --- | --- | --- | --- | --- | --- | --- | --- | --- | --- |
| **VAMC** | 20 | 74.7<br>(6.0) | 1.76<br>(0.06) | 96.9<br>(13.5) | 9.8<br>(2.4) | 14.8<br>(4.4) | 0.80 (0.16) | 1.2 (0.23) | 14.7<br>(6.3) | 25.0 (7.3) |
| **Baylor** | 18 | 75.1<br>(11.7) | 1.74<br>(0.08) | 80.9<br>(18.1) | 8.2<br>(3.0) | 15.1<br>(4.2) | 0.87 (0.25) | 1.28 (0.39) | 13.2<br>(5.2) | 28.1 (8.2) |
| **M Health**<br>**Fairview** | 9 | 70.1<br>(7.8) | 1.79<br>(0.12) | 87.6<br>(10.5) | 6.6<br>(4.1) | 15.0<br>(3.2) | 1.02 (0.28) | 1.32 (0.27) | 10.8<br>(2.3) | 23.6 (5.2) |
| **Harvard** | 5 | 78.2<br>(5.1) | 1.71<br>(0.1) | 95.0<br>(22.0) | 5.2<br>(2.9) | 14.0<br>(3.4) | 0.77 (0.25) | 1.07 (0.45) | 15.0<br>(3.4) | 28.3 (6.6) |

Datasets from the current study are available upon request. The raw data supporting the conclusions of this article will be made available to qualified researchers, without undue reservation.

## 3 Results

### 3.1 Enrollment and Allocation

The flow chart for the study is shown in Figure 2. Sixty-seven subjects were assessed for eligibility across the four participating sites. Their respective baseline characteristics are shown in Table 1. First enrollment occurred at the MVAHCS on 10/22/2018, followed by M Health Fairview/University of Minnesota on 10/23/2018, Baylor College of Medicine on 12/20/2018 and Marcus Institute/Harvard Medical School on 09/19/2019. The last enrollment occurred at Baylor College of Medicine on 01/10/2020. Subjects across the four sites were pooled for the continued analysis presented here.

**Figure 2.**
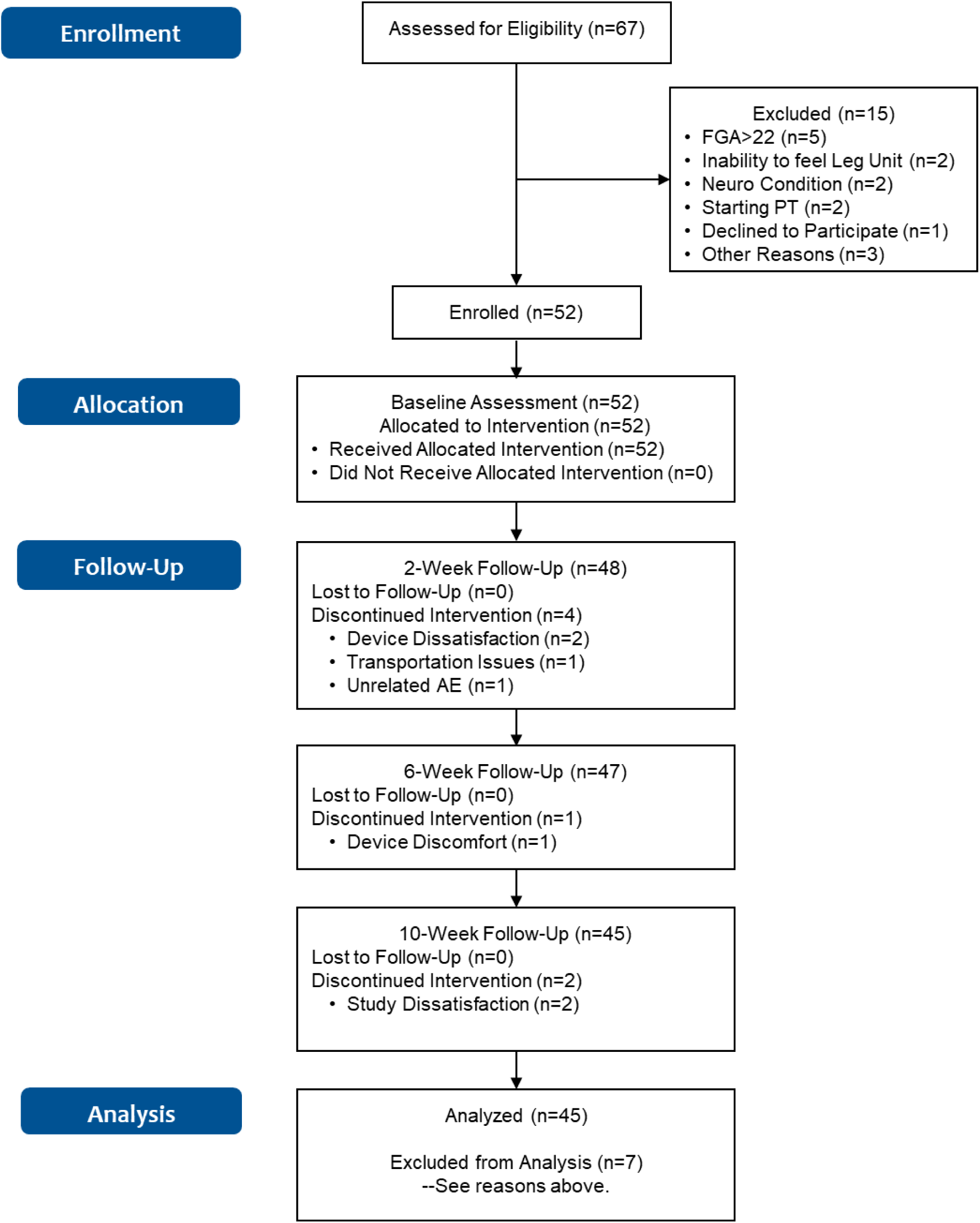
Flowchart of the study.

Fifteen subjects were excluded from participation of which five had an FGA score higher than 22; two were unable to sense tactile stimuli from the device; two had neurological conditions that prevented device use; two were planning to start physical therapy treatment, one declined to participate and three for other reasons. Fifty-two subjects were enrolled for baseline assessment and allocated for the intervention. A total of seven subjects discontinued participation, four subjects at the two-week assessment (two related to device use, one due to transportation issues, and one due to an unrelated adverse event, Figure 2), one ahead of the six-week assessment and two ahead of the 10- week assessment (due to device and study issues), respectively (Figure 2). A total of 45 subjects (87%) completed all in-clinic assessments from baseline to the 10-week endpoint.

### 3.2 Baseline Characteristics and Outcomes

Table 2 shows baseline characteristics of all enrolled participants (n=52), then separately for subjects who reported having fallen in the six months preceding the study (Pre-Faller, n=30) and those who did not report a fall (Pre-NonFaller, n=22). Overall, participants were elderly (74.4±8.7 yrs.), overweight (BMI >25) and mostly male (79%). A majority used an assistive device (54%) and had fallen in the previous year (71%) or in the past six months (58%). Furthermore, a majority showed high fall risk based on low 4-Stage Balance Test outcomes (63% of subjects < 30s) (CDC, 2017) or low ABC scores (56% of subjects scored < 67%) (Lajoie and Gallagher, 2004). Twenty-five percent of participants had a normal gait speed less than 0.7 m/s (Montero-Odasso et al., 2005) and half of the participants performed the TUG slower than 12 s (CDC, 2017), the commonly used thresholds for high fall risk. The Pre-Faller group had a higher number of fall-risk factors as compared to the Pre- NonFaller group (5.3±1.0, vs. 3.5±1.3, respectively, p<0.000001). The baseline FGA score was statistically significantly lower in Pre-Faller (13.5±3.7) as compared to Pre-NonFaller (16.7±3.6, p<0.004). There was no difference in normal gait speed between the two groups (p=0.12), although fast gait speed was higher in the Pre-NonFaller (1.41±0.35 m/s) as compared to the Pre-Faller subjects (1.13±0.34 m/s, p<0.006). Furthermore, TUG times were significantly slower in the Pre- Faller group compared to the Pre-NonFaller (14.7±6.3s and 12.0±2.9, respectively, p<0.05).

**Table 2.** Baseline characteristics of subjects enrolled in the study (n=52), then separately for subjects who reported having fallen in the past six months (Pre-F) and those who did not (Pre-NF). Values shown are Mean (Standard Deviation). Column p-level shows significance level for comparison between the Pre-F and Pre-NF groups. In bold if p<0.05.

| Baseline Assessment | All<br>n=52 | Pre-F<br>n=30 | Pre-NF<br>n=22 | p-level |
| --- | --- | --- | --- | --- |
| Gender Female (n) | 11 of 52 (21%) | 9 of 30 (30%) | 2 of 22 (10%) | 0.069 |
| Use of Assistive Device (n) | 28 of 52 (54%) | 19 of 30 (63%) | 9 of 22 (41%) | 0.11 |
| Gait Speed Normal <0.7m/s (n) | 13 of 52 (25%) | 10 of 30 (33%) | 3 of 22 (14%) | 0.10 |
| Timed Up and Go >12s (n) | 26 of 52 (50%) | 18 of 30 (60%) | 8 of 22 (36%) | 0.09 |
| 4-Stage Balance Test <30s (n) | 33 of 52 (63%) | 22 of 30 (73%) | 11 of 22 (50%) | 0.08 |
| ABC Score <67% (n) | 30 of 52 (56%) | 20 of 30 (67%) | 10 of 22 (45%) | 0.13 |
| Fallen in Last 6 Months (n) | 30 of 52 (58%) | 30 of 30 (100%) | 0 | n/a |
| Fallen in Last 12 Months (n) | 37 of 52 (71%) | 30 of 30 (100%) | 7 of 22 (32%) | **<0.00001** |
| Number of Falls 6 Months | 65 | 65 | 0 | n/a |
| Number of Falls 12 Months | 121 | 109 of 121 (90%) | 12 of 121 (10%) | **<0.000001** |
|  | Mean (SD)<br>n=52 | Mean (SD)<br>n=30 | Mean (SD)<br>n=22 | p-level |
| Age (yrs) | 74.4 (8.7) | 73.8 (9.1) | 75.2 (8.2) | 0.57 |
| Height (m) | 1.75 (0.08) | 1.76 (0.09) | 1.75 (0.07) | 0.54 |
| Weight (kg) | 89.6 (16.8) | 89.7 (17.8) | 89.3 (15.8) | 0.93 |
| BMI (kg/m<sup>2</sup>) | 29.1 (5.2) | 29.0 (5.8) | 29.2 (5.2) | 0.92 |
| FGA Score | 14.9 (4.0) | 13.5 (3.7) | 16.7 (3.6) | **<0.004** |
| Gait Speed, Normal (m/s) | 0.86 (0.23) | 0.81 (0.26) | 0.92 (0.18) | 0.12 |
| Gait Speed, Fast (m/s) | 1.25 (0.37) | 1.13 (0.34) | 1.41 (0.35) | **<0.006** |
| TUG (s) | 13.5 (5.3) | 14.7 (6.3) | 12.0 (2.9) | **<0.05** |
| 4-Stage Balance Test (s) | 26.2 (7.3) | 24.8 (6.3) | 28.0 (8.3) | 0.055 |
| Fall-Risk Factors* (n of 7) | 4.5 (1.5) | 5.3 (1.0) | 3.5 (1.3) | **<0.000001** |
| ABC Score (%) | 60.8 (17.6) | 57.0 (15.2) | 66.0 (19.7) | 0.07 |
| VADL Mean Score | 3.66 (1.07) | 3.94 (1.04) | 3.29 (1.02) | **<0.03** |
| VAS Pain Score (0-10) | 2.7 (2.2) | 2.6 (2.1) | 2.9 (2.4) | 0.67 |
| PHQ-9 | 4.4 (3.8) | 5.2 (4.3) | 3.4 (3.8) | 0.11 |
| Pain Interference<br>PROMIS® 6b | 51.1 (8.0) | 52.5 (8.1) | 49.0 (8.1) | 0.10 |
| Satisfaction Social Roles<br>PROMIS® 8a | 50.4 (7.8) | 49.1 (7.7) | 52.1 (7.8) | 0.13 |
| Ability to Participate<br>PROMIS® 8a | 50.0 (7.2) | 49.8 (7.1) | 50.2 (7.2) | 0.88 |

\*Fall-risk factors assessed in the current study included, recent history of falls (Tinetti and Kumar, 2010), PN diagnosis (Richardson and Hurvitz, 1995), FGA score<23 (Wrisley and Kumar, 2010), TUG >12 s (CDC, 2017), 4-Stage Balance Test<30s (CDC, 2017), Gait Speed <0.7m/s (Studenski et al., 2003; Montero-Odasso et al., 2005), ABC score <67% (Lajoie and Gallagher, 2004).

The ABC score was higher in the Pre-NonFaller compared to the Pre-Faller group, although the difference was not statistically significant (66.0±19.7 vs. 57.0±15.2, respectively, p=0.07). The VADL score was marginally lower in the Pre-NonFaller group (3.29±1.02, vs. 3.94±1.04, p<0.03). Pain scores were similar for the two groups (2.6±2.1 vs. 2.9±2.4). The PHQ-9 score was slightly higher in the Pre-Faller group, although the difference was not statistically significant (p=0.11). PROMIS outcome scores for “Pain Interference,” “Satisfaction with Social Roles,” and “Ability to Participate” (Table 2) showed mean values around 50 for both groups, which is considered average for the US population (Askew et al., 2016; Hahn et al., 2016a; Hahn et al., 2016b). Any differences were well within 10, which is one standard deviation of these measures in the US population (Table 2) (Askew et al., 2016; Hahn et al., 2016a; Hahn et al., 2016b).

### 3.3 Chronic Conditions and Medication Use

Table 3 shows self-reported chronic conditions and medication use for subjects enrolled in the study. On average, subjects reported having 8.2±3.3 chronic conditions. All subjects had a diagnosis of PN either in their medical chart or provided in a letter signed by their physician. Most subjects reported having neuropathic pain in their feet (73%) as well hypertension (63%) and half of participants reported having chronic back pain. All subjects reported having difficulty with walking and balance. The Pre-Faller group reported a higher incidence of cancer as a chronic condition than the Pre- NonFaller group (43% vs. 14%, respectively, p<0.03). Falls in the 12 months preceding study participation were reported by all the Pre-Faller participants (i.e. reporting falls over the past 12 and six months) versus by seven of the 22 in the Pre-NonFaller group (i.e. reporting falls over the past 12 but not six months) (p<0.00001). Ninety percent of the falls reported in the 7-12 months preceding the study were reported by the Pre-Faller participants (p<0.000001). Overall, participants reported taking one non-prescription medication and eight (median) prescription medications of which three are known to cause potential balance issues and increase the risk of falling (Woolcott et al., 2009). Medication use was similar between the Pre-Faller and Pre-NonFaller groups (Table 3).

**Table 3.** Self-reported chronic conditions as well as medication use for subjects enrolled in the study. Column p-level shows significance level for comparison between the Pre-F and Pre-NF groups. In bold if p<0.05.

| Baseline Assessment | All<br>n=52 | Pre-F<br>n=30 | Pre-NF<br>n=22 | p-level |
| --- | --- | --- | --- | --- |
| Number of Chronic Conditions (n) | 8.2 (3.3) | 8.6 (3.2) | 7.8 (3.4) | 0.39 |
| Peripheral Neuropathy (n) | 52 (100%) | 30 (100%) | 22 (100%) | n/a |
| Numbness in Feet (n) | 49 (94%) | 30 (100%) | 19 (86%) | **<0.05** |
| Neuropathic Pain in Feet (n) | 38 (73%) | 22 (73%) | 16 (73%) | 0.96 |
| Hypertension (n) | 33 (63%) | 19 (63%) | 14 (64%) | 0.98 |
| Back Pain (n) | 26 (50%) | 16 (53%) | 10 (45%) | 0.57 |
| Arthritis (n) | 24 (46%) | 15 (50%) | 9 (41%) | 0.52 |
| Knee Dysfunction (n) | 23 (44%) | 14 (47%) | 9 (41%) | 0.68 |
| Diabetes Diagnosis (n) | 19 (37%) | 9 (30%) | 10 (45%) | 0.25 |
| Inner Ear Problems (n) | 17 (33%) | 9 (30%) | 8 (36%) | 0.63 |
| Heart Disease (n) | 16 (31%) | 10 (33%) | 6 (27%) | 0.64 |
| Neck Pain (n) | 16 (31%) | 9 (30%) | 7 (32%) | 0.89 |
| Cancer (n) | 16 (31%) | 13 (43%) | 3 (14%) | **<0.03** |
| Lung Disease (n) | 10 (19%) | 7 (23%) | 3 (14%) | 0.38 |
| Stroke (n) | 9 (17%) | 4 (13%) | 5 (23%) | 0.38 |
| Osteoporosis(n) | 10 (19%) | 7 (23%) | 3 (14%) | 0.38 |
| Seizures (n) | 5 (10%) | 3 (10%) | 2 (9%) | 0.91 |
| Ankle Dysfunction (n) | 5 (10%) | 4 (13%) | 1 (5%) | 0.29 |
| TMJ/Jaw Pain (n) | 5 (10%) | 2 (7%) | 3 (14%) | 0.40 |
| Fainting (n) | 5 (10%) | 2 (7%) | 3 (14%) | 0.40 |
| Migraines (n) | 4 (8%) | 2 (7%) | 2 (9%) | 0.75 |
| Meningitis (n) | 0 | 0 | 0 | n/a |
| Other Conditions (n) | 14 (27%) | 11 (37%) | 3 (14%) | 0.06 |
| Difficulty Walking/balance (n) | 52 (100%) | 30 (100%) | 22 (100%) | n/a |
|  | **Mean (SD)**<br>**Median** | **Mean (SD)** | **Mean (SD)** | **p-level** |
| Prescription Medications (n) | 8 | 8.5 | 7 | 0.50 |
| Non-Prescription Medications (n) | 1 | 1 | 1 | n/a |
| Medications Associated with Falls (n) | 3 | 3 | 3 | n/a |

### 3.4 Clinical Outcomes

Table 4 shows clinical outcomes for the 45 subjects who completed assessments at baseline, 2, 6 and 10 weeks. For all subjects, repeated measures ANOVA showed highly statistically significant differences across the assessment events for all subjects and all clinical outcomes (0.00001 < p < 0.01) except for the 4-Stage Balance Test, which was not statistically significant (p=0.23, Table 4 ANOVA column). Further pairwise comparisons following statistically significant ANOVA showed highly statistically significant differences between the baseline assessment and the 2, 6, and 10-week assessments, respectively, for the FGA score and normal gait speed and between baseline and 6 and 10 weeks, respectively, for fast gait speed and the TUG scores. Cohen’s d effect size calculated between the baseline and 10-week endpoint was large for FGA (0.92, FGA change from 15.0 to 19.1) and small to medium for normal gait speed (0.42, 0.86m/s to 0.95m/s), fast gait speed (0.27, 1.24 m/s to 1.33 m/s) and the TUG (0.28, 13.8 s to 12.5 s, Table 4).

**Table 4.** Clinical outcomes for the 45 subjects completing all assessments for baseline, 2, 6 and 10 weeks as well as the subgroups of Pre-Fallers and Pre-NonFallers. Values in **bold** indicate statistical significance. ANOVA column shows significance levels of main effect from initial repeated measures ANOVA test. If significant (p<0.05), pairwise comparisons were made using three dependent t-tests between baseline and 2, 6 and 10-week assessments, respectively. Bonferroni’s adjustment of significance levels for correlated measures was applied. As noted, values in *(italics)* indicate Pearson’s correlation coefficient followed by the adjusted significance level required for an overall significance of 0.05 as marked with *. If ANOVA was non-significant, no further comparisons were made. Cohen’s d indicates effect size where 0.2 is represents a small effect, 0.5 a medium effect, and 0.8 a large effect. Values in parenthesis show 95% confidence interval.

| **ALL** | **Baseline**<br>**Mean (SD)**<br>**n=45** | **2-week**<br>**Mean (SD)**<br>**n=45** | **6-week**<br>**Mean (SD)**<br>**n=45** | **10-week**<br>**Mean (SD)**<br>**n=45** | **ANOVA**<br>**p-level** | **Cohen's d**<br>**(95% CI's)** |
| --- | --- | --- | --- | --- | --- | --- |
| FGA Score | 15.0 (4.0) | 18.3 (4.4)<br>**<0.0000001** | 18.5(4.4)<br>**<0.0000001** | 19.1 (5.2)<br>**<0.0000001** | **<0.00001** | 0.92<br>(0.49, 1.35) |
| Gait Speed<br>Normal (m/s) | 0.86 (0.24) | 0.92 (0.26)<br>**<0.002** | 0.94 (0.25)<br>**<0.0001** | 0.95 (0.24)<br>**<0.00005** | **<0.00001** | 0.42<br>(0.01, 0.82) |
| Gait Speed Fast<br>(m/s) | 1.24 (0.37) | 1.27 (0.33)<br>0.30 | 1.30 (0.37)<br>**<0.016** | 1.33 (0.38)<br>**<0.00168** | **<0.013** | 0.27<br>(-0.14,0.67) |
| TUG (s) | 13.8 (5.5) | 12.7 (4.2)<br>0.06 | 12.3 (4.2)<br>**<0.034 (0.69)**<br>0.0397* | 12.5 (3.7)<br>**<0.012** | **<0.01** | 0.28<br>(-0.12, 0.68) |
| 4-Stage Balance<br>Test (s) | 26.2 (7.1) | 27.1 (7.7) | 27.7 (7.7) | 27.8 (6.8) | 0.23 | n/a |
| **Pre-Fallers** | **Baseline**<br>**Mean (SD)**<br>**n=25** | **2-week**<br>**Mean (SD)**<br>**n=25** | **6-week**<br>**Mean (SD)**<br>**n=25** | **10-week**<br>**Mean (SD)**<br>**n=25** | **p-level**<br>**ANOVA** | **Cohen's d**<br>**(95% CI's)** |
| FGA Score | 13.5 (3.6) | 16.6 (3.7)<br>**<0.0000005** | 17.3 (4.1)<br>**<0.00002** | 16.8 (4.9)<br>**<0.00016** | **<0.00001** | 0.82<br>(0.26, 1.39) |
| Gait Speed<br>Normal (m/s) | 0.79 (0.27) | 0.85 (0.26)<br>**<0.022 (0.90)**<br>0.047* | 0.89 (0.25)<br>**<0.0021** | 0.86 (0.24)<br>**<0.016** | **<0.0014** | 0.28<br>(-0.27, 0.83) |
| Gait Speed Fast<br>(m/s) | 1.09 (0.33) | 1.16 (0.30)<br>**<0.035 (0.88)**<br>0.046* | 1.17 (0.32)<br>**<0.0165** | 1.17 (0.35)<br>**<0.015** | **<0.028** | 0.27<br>(-0.28, 0.82) |
| TUG (s) | 15.3 (6.6) | 14.0 (4.6)<br>0.14 | 13.3 (3.8)<br>0.047 | 13.7 (4.2)<br>0.056 | **<0.025** | n/a |
| 4-Stage Balance<br>Test (s) | 24.7 (5.5) | 25.4 (7.8) | 26.0 (7.9) | 27.5 (6.1) | 0.15 | n/a |
| **Pre-NonFallers** | **Baseline**<br>**Mean (SD)**<br>**n=20** | **2-week**<br>**Mean (SD)**<br>**n=20** | **6-week**<br>**Mean (SD)**<br>**n=20** | **10-week**<br>**Mean (SD)**<br>**n=20** | **p-level**<br>**ANOVA** | **Cohen's d**<br>**(95% CI's)** |
| FGA Score | 16.9 (3.6) | 20.5 (4.4)<br>**<0.00009** | 20.1 (4.3)<br>**<0.00004** | 22.0 (4.0)<br>**<0.000004** | **<0.0000001** | 1.38<br>(0.70, 2.1) |
| Gait Speed<br>Normal (m/s) | 0.93 (0.17) | 1.01 (0.22)<br>**<0.031 (0.78)**<br>0.043* | 1.01 (0.24)<br>**<0.019 (0.84)**<br>0.045* | 1.06 (0.20)<br>**<0.0004** | **<0.0011** | 0.74<br>(0.10, 1.37) |
| Gait Speed Fast<br>(m/s) | 1.41 (0.35) | 1.42 (0.33) | 1.43 (0.41) | 1.52 (0.34) | 0.10 | n/a |
| TUG (s) | 11.9 (2.9) | 11.2 (3.2) | 11.7 (5.0) | 10.9 (2.2) | 0.49 | n/a |
| 4-Stage Balance<br>Test (s) | 27.6 (8.5) | 29.4 (7.1) | 29.9 (7.1) | 28.1 (7.7) | 0.45 | n/a |

Both the Pre-Faller and Pre-NonFaller groups increased their FGA scores from baseline to the 2-, 6-, and 10-week assessments, respectively (p<0.00016). The Pre-Faller group had lower FGA scores both at baseline as compared to the Pre-NonFaller (13.5 vs. 16.9) and at the 10-week assessment (16.8 vs. 22.0). Effect sizes for FGA changes for both groups were large, although higher in the Pre- NonFaller group compared to the Pre-Faller group (Cohen’s d 1.38 vs 0.82, respectively). Similarly, normal gait speed increased for both groups although the effect size was larger in the Pre-NonFaller group (Cohen’s d 0.74 vs. 0.28, respectively). Interestingly, fast gait speed only improved in the Pre- Faller group from 1.09 m/s at baseline to 1.17 m/s at 10 weeks (p <<u>0.015, Table 4</u>) although effect size was small (Cohen’s d 0.27). There was an overall statistically significant change in TUG for the Pre-Faller group (ANOVA, p<0.025) although none of the pairwise comparisons reached statistical significance required after Bonferroni correction (0.047 < p <0.14). The overall ANOVA for TUG in the Pre-NonFaller group was not statistically significant (p=0.49, Table 4).

### 3.5 Participant Reported Outcomes, Sensation Tests, and Device Use

Patient reported outcomes for the 45 subjects who completed all assessments are shown in Table 5 as well as for the Pre-Faller and Pre-NonFaller groups. For all subjects, there was an overall significant ANOVA for the VADL score (p<0.044) although none of the pairwise comparisons reached statistical significance following Bonferroni correction (0.053 < p <0.99). There were no other statistically significant differences across all subjects during the 10-week period.

**Table 5.** Results from participant-reported outcomes and Rydel-Seiffer vibration sensation screening for the 45 subjects who completed all assessments. Data is shown separately for the group as a whole and for Pre-Fallers (having reported fallen in the previous 6 months) and Pre-NonFallers (no falls reported in the previous 6 months). Values in *(italics)* indicate Pearson’s correlation coefficient followed by the Bonferroni adjusted significance level required for an overall significance of 0.05 as marked with *. There were no differences in vibration sensation between Pre-Fallers and Pre- NonFallers and no differences between baseline and 10-week assessments.

| **ALL (n=45)** | **Baseline**<br>**Mean (SD)**<br>**n=45** | **2-week**<br>**Mean (SD)**<br>**n=45** | **6-week**<br>**Mean (SD)**<br>**n=45** | **10-week**<br>**Mean (SD)**<br>**n=45** | **ANOVA**<br>**p-level** |
| --- | --- | --- | --- | --- | --- |
| ABC-Score (%) | 61.4 (17.9) | 65.0 (13.1) | 64.2 (15.1) | 65.1 (14.1) | 0.36 |
| VADL Mean Score | 3.70 (1.09) | 3.37 (0.86)<br>0.053 | 3.52 (0.99)<br>0.16 | 3.63 (0.93)<br>0.99 | **<0.044** |
| VAS Score (0-10) | 2.8 (2.2) | 2.5 (2.1) | 2.5 (2.2) | 2.6 (2.3) | 0.31 |
| PHQ-9 | 4.5 (3.9) | 3.8 (3.5) | 3.6 (3.8) | 3.9 (4.5) | 0.25 |
| Pain Interference<br>PROMIS® 6b | 50.8 (7.9) | 51.2 (8.1) | 50.5 (8.9) | 51.8 (8.5) | 0.85 |
| Satisfaction Social Roles<br>PROMIS® 8a | 50.2 (7.8) | 52.1 (7.2) | 51.7 (7.3) | 52.1 (7.8) | 0.10 |
| Ability to Participate<br>PROMIS® 8a | 49.8 (7.3) | 50.7 (6.4) | 51.0 (7.1) | 51.3 (8.1) | 0.38 |
| R 1st MTP Joint | 2.0 (2.5) | n/a | n/a | 2.0 (2.5) | n/a |
| R Lateral Malleolus | 3.6 (2.7) | n/a | n/a | 3.4 (2.6) | n/a |
| R Patella | 4.5 (2.0) | n/a | n/a | 4.5 (1.8) | n/a |
| L 1st MTP Joint | 2.3 (2.6) | n/a | n/a | 2.2 (2.7) | n/a |
| L Lateral Malleolus | 3.7 (2.7) | n/a | n/a | 3.6 (2.5) | n/a |
| L Patella | 4.3 (2.1) | n/a | n/a | 4.4 (1.6) | n/a |
| **Pre-Fallers (n=25)** | **Baseline**<br>**Mean (SD)** | **2-week**<br>**Mean (SD)** | **6-week**<br>**Mean (SD)** | **10-week**<br>**Mean (SD)** | **p-level** |
| ABC-Score (%) | 57.7 (17.9) | 61.5 (14.0) | 61.8 (15.1) | 61.2 (13.9) | 0.29 |
| VADL Mean Score | 4.00 (1.13) | 3.68 (0.90) | 3.83 (1.01) | 3.91 (0.97) | 0.42 |
| VAS Score (0-10) | 2.5 (2.4) | 2.5 (2.5) | 2.8 (2.6) | 2.6 (2.5) | 0.55 |
| PHQ-9 | 5.3 (3.9) | 4.7 (3.3) | 3.9 (3.3) | 4.5 (3.5) | 0.08 |
| Pain Interference<br>PROMIS® 6b | 51.9 (8.3) | 52.2 (8.5) | 51.6 (9.9) | 53.6 (8.0) | 0.90 |
| Satisfaction Social Roles<br>PROMIS® 8a | 49.1 (8.0) | 49.9 (7.9) | 49.2 (7.5) | 49.6 (7.7) | 0.68 |
| Ability to Participate<br>PROMIS® 8a | 49.9 (7.5) | 48.6 (5.8) | 49.5 (6.8) | 49.4 (8.0) | 0.58 |
| **Pre-NonFallers (n=20)** | **Baseline**<br>**Mean (SD)** | **2-week**<br>**Mean (SD)** | **6-week**<br>**Mean (SD)** | **10-week**<br>**Mean (SD)** | **p-level** |
| ABC-Score (%) | 66.2 (18.0) | 69.8 (11.9) | 67.2 (15.1) | 70.2 (14.6) | 0.96 |
| VADL Mean Score | 3.32 (1.06) | 2.94 (0.80) | 3.11 (0.99) | 3.28 (0.89) | 0.08 |
| VAS Score (0-10) | 3.0 (2.0) | 2.4 (1.5) | 2.2 (1.7) | 2.7 (2.0) | 0.10 |
| PHQ-9 | 3.5 (3.9) | 2.6 (3.7) | 3.2 (4.4) | 3.2 (5.6) | 0.66 |
| Pain Interference<br>PROMIS® 6b | 49.4 (7.6) | 49.8 (7.6) | 49.1 (7.6) | 49.3 (9.1) | 0.91 |
| Satisfaction Social Roles<br>PROMIS® 8a | 51.7 (7.8) | 55.1 (6.4) | 54.8 (7.0) | 55.4 (8.0) | 0.12 |
| Ability to Participate<br>PROMIS® 8a | 49.7 (7.1) | 53.7 (7.3)<br>**<0.02** (0.64)<br>0.038* | 52.8 (7.6)<br>0.06 | 53.7 (8.4)<br>0.05 | **<0.041** |

Table 5 shows and data from vibration sensation testing using the Rydel-Seiffer graduated tuning fork for all subjects. There were no differences in vibration sensation between Pre-Faller and Pre- NonFaller and no differences between baseline and 10-week assessments. Vibration sensitivity showed bilateral symmetry across the three sites tested, right and left patella, lateral malleolus and first metatarsophalangeal joints, respectively. There was a gradual proximal to distal decrease in vibration sensitivity across the three sites from 4.3-4.4 proximally and 2.0 distally based on the 0-8 Rydel-Seiffer scale (Table 5). Similarly, there was bilateral symmetry for the WEST monofilament sensitivity test. At baseline, the median for the monofilament test was 50g for both feet and across all four anatomical test sites, the first, third, and fifth metatarsal heads as well as the great toe. This was similar at the 10-week assessment with the exception for the first metatarsals of both feet where the median monofilament increased to 200g.

In the Pre-Faller subgroup, there were no overall statistically significant differences for any of the participant reported outcomes across the 10-week period (Table 5). For the Pre-NonFaller group, there was a statistically significant difference in the PROMIS “Ability to Participate” score at two weeks (ANOVA, p<0.04, 49.7±7.1 to 53.7±7.3, p<0.02, Table 5). The improvement appeared maintained at six (52.8±7.6) and 10 weeks (53.7±8.4) although it did not fully reach statistical significance (p<0.06). None of the other participant reported outcomes for the Pre-NonFaller group were statistically different across the 10-week period (Table 5). Subjects reported using the device on average 5.5±1.4 days/week (range 3.5-7.0) for a total of 36.1±14.9 hours/week (range 5.3-56.0).

### 3.6 Falls Assessment

Parameters related to falls and fall risk assessed at baseline and throughout the 10 weeks are shown in Table 6. Overall, after 10 weeks, 13 of the 45 subjects had achieved FGA scores higher than 22, the cut-off for normal fall risk (Wrisley and Kumar, 2010). Four of these subjects were part of the Pre-Faller subgroup and nine were from the Pre-NonFaller group. Subjects reported a total of 62 falls in the six months prior to the study. During the 10-week study period, 17 falls were reported, 13 of which occurred in the Pre-Faller group and four in the Pre-NonFaller group (Table 6). Sixteen of the 25 Pre-Fallers did not fall during the 10-week period of the trial. There was a non-significant decrease in fall rate across all subjects (from 7.7 to 5.4 falls/1000 patient days, p<0.27). In the Pre- Faller group, there was a 46% statistically significant decrease in fall rate (from 13.8 to 7.4 falls per 1000 patient days, p<0.014). The Pre-NonFaller group showed a smaller non-significant increase in fall rate (from 0 to 2.9 falls per 1000 patient days, p=0.125). This increase was based on one fall each by four subjects who had not fallen in the prior six months (Table 6). There was a statistically significant decrease in the number of fall risk factors (Table 6) from baseline to 10 weeks across all subjects (from 4.2 to 3.8 fall risk factors, p<0.047). This decrease was larger and statistically significant in the Pre-Faller group (from 5.1 to 4.3 fall risk factors, p<0.023). There was no change in the number of fall risk factors in the Pre-NonFaller group (p=0.76, Table 6).

**Table 6.** Parameters related to falls and fall risk assessed at baseline and at 10 weeks. Fall rates are reported in number of falls per 1000 patient days. Fall risk factors are identified in Table 2. *Wilcoxon Signed Rank test

| **All (n=45)** | **Baseline** | **10 Weeks** | **p-level** |
| --- | --- | --- | --- |
| Number of Subjects FGA>22 | 0 of 45 | 13 of 45 | n/a |
| #Falls (pre-6 mo & in study) | 62 | 17 | n/a |
| Fall Rate (pre-6mo & in study) | 7.7 | 5.4 | 0.27* |
| #Fallers (pre-6mo & in study) | 25 | 22 | n/a |
| # Fall Risk Factors | 4.2 (1.5) | 3.8 (1.6) | **<0.047** |
| **Pre-Fallers (n=25)** |  |  |  |
| Number of Subjects FGA>22 | 0 of 25 | 4 of 25 | n/a |
| #Falls (pre-6 mo & in study) | 62 | 13 | n/a |
| Fall Rate (pre-6mo & in study) | 13.8 | 7.4 | **<0.014\*** |
| #Fallers (pre-6mo & in study) | 25 | 9 | n/a |
| # Fall Risk Factors | 5.1 (1.3) | 4.3 (1.7) | **<0.023** |
| **Pre-NonFallers (n=20)** |  |  |  |
| Number of Subjects FGA>22 | 0 of 20 | 9 of 20 | n/a |
| #Falls (pre-6 mo & in study) | 0 | 4 | n/a |
| Fall Rate (pre-6mo & in study) | 0 | 2.9 | 0.125* |
| #Fallers (pre-6mo & in study) | 0 | 4 | n/a |
| # Fall Risk Factors | 3.2 (1.8) | 3.1 (1.7) | 0.76 |

## 4 Discussion

### 4.1 Key Findings

Results from this multi-site clinical trial supported our *a priori* hypothesis that patients with gait and balance problems due to sensory PN and high risk of falls would show clinically meaningful improvements of gait and dynamic balance function after 10 weeks of using a wearable sensory neuroprosthesis, confirming previously demonstrated in-clinic findings in a randomized controlled cross-over trial (Koehler-McNicholas et al., 2019). Overall, the mean FGA score improved from 15.0 at baseline to 19.1 at 10 weeks across all subjects (Table 4), beyond the MCID for the FGA (Beninato et al., 2014). Thirteen subjects reached normal fall risk status showing an FGA score higher than 22 (Wrisley and Kumar, 2010) after 10 weeks of device use. Both normal and fast gait speed improved overall by 0.09m/s, near what is considered substantial change (0.10m/s) for older adults (Perera et al., 2006). Across both groups, TUG improved from 13.8 s to 12.5 s, which is beyond the MDC for older adults with type 2 diabetes (Alfonso-Rosa et al., 2014). Effect sizes ranged from large for FGA scores (Cohen’s d, 0.92) to small for fast gait speed and TUG (0.27 and 0.28, respectively).

As expected, vibratory sensation did not improve during the trial suggesting new sensory information from the device provided relevant input to improve function. Although both Pre-NonFallers and Pre- Fallers showed improvements, effect sizes were larger in the Pre-NonFaller group for FGA and normal gait speed while only the Pre-Faller group showed a statistically significant improvement in fast gait speed with an effect size of 0.27. The increase in fast gait speed from 1.41 m/s to 1.52 m/s in the Pre-NonFaller group did not reach statistical significance (ANOVA main effect, p=0.10), although such gait speeds are considered in the high range of elderly community ambulators (Middleton et al., 2015). Interestingly, the 4-Stage Balance measure, an indicator of static balance, did not improve significantly over the 10-week period in either of the two subgroups although our recent in-clinic study (Koehler-McNicholas et al., 2019) found a statistically significant improvement in this measure in a group of PN patients. One important difference between these two groups may be a slightly lower baseline static balance performance in the first cohort, a mean of 22.2 s versus 26.2 s in the current one. The first group improved their performance in clinic to a mean of 27.6 s (p<0.001) while they improved to a nearly identical 27.8 s (n.s.) in the current study. It may be that further improvement of static balance performance would require some additional training challenge in addition to device use (e.g. Romberg and sharpened Romberg types of activities) and simply wearing the device daily does not sufficiently challenge the static balance ability assessed by the 4-Stage Balance Test.

### 4.2 Physical Intervention versus Sensory Substitution

To our knowledge, this is the first trial where a cohort of subjects with gait and balance problems related to PN have worn a sensory prosthetic device of this kind in the community and on a regular basis. Other wearable neuromodulation technologies have been used as a treatment modality in a home setting, although typically only worn in conjunction with a home therapy program for balance and mobility in different categories of subjects, e.g., multiple sclerosis (Leonard et al., 2017) and healthy elderly (Bao et al., 2018). A recent case study of a patient with PN wearing Walkasins for a year (Wrisley et al., 2018), found dramatic improvements in gait and balance outcomes when daily continuous device use was combined with balance therapy. The patient had received balance therapy twice a week for over five months prior to using Walkasins and no longer noticed any further improvements (Wrisley et al., 2018). In the current study, in an attempt to isolate the effect of long-term device use on clinical outcomes, subjects were instructed to wear the device as much as possible and were not allowed to participate in any additional balance training or therapy. Furthermore, they were not systematically informed of any changes in their outcomes or performance, which they would normally receive in regular clinical care. Still, improvements in clinical outcomes were similar to those reported after various balance intervention programs conducted over comparable time periods up to 12 weeks in similar patient populations (Shumway-Cook et al., 1997; Wolf et al., 2001; Li et al., 2005; Manor et al., 2014), or even longer up to six months and nearly a year (Wolf et al., 2003; Li et al., 2005; Li and Manor, 2010).

Interestingly, two systematic reviews of interventions specifically for patients with diabetic PN gave lower extremity strengthening only a fair recommendation while other interventions showed insufficient evidence to increase function (Ites et al., 2011; Tofthagen et al., 2012). More recently, improvement in function in patients with PN following specific task-oriented training has been shown (Salsabili et al., 2015). Consequently, it may be particularly difficult to enhance gait and balance function in individuals with PN using physical therapy or balance training activities alone, possibly because these interventions do not replace lost somatosensory input. Furthermore, effects of a training or therapy program on gait and balance function are likely due to different mechanisms than the use of a sensory substitution device.

For training/therapy programs to be effective, well known principles of training and exercise physiology must be adhered to (Oddsson et al., 2007) ensuring that sensorimotor systems are sufficiently challenged to adapt and improve their capabilities leading to improved muscle function and neuromotor coordination. However, patients with PN have lost important cutaneous afferent systems that typically would not be affected by such interventions. Instead, improvements related to use of Walkasins seen in the current study are most likely due to participants receiving new tactile balance stimuli that are relevant for gait and balance and therefore become integrated into their neuromotor control and movement repertoire. Subjects in the current study received hundreds of such stimuli per hour from the device during their regular standing and walking activities throughout the day.

Consequently, we hypothesize that any balance-related therapy or training activity in conjunction with wearing the device would provide an additive effect to overall function and balance outcomes. This hypothesis is supported by our recent case study (Wrisley et al., 2018) as well as our previous in-clinic study where subjects following a baseline assessment were randomized to either wearing the device turned on or turned off while performing a brief 10-15 min standardized balance activity session with a physical therapist (Koehler-McNicholas et al., 2019). Ten of 15 subjects in the on group increased their FGA scores by at least 4 points compared to five of 16 in the off group (p<0.05). Furthermore, seven of 15 subjects in the on group increased gait speed by >0.13m/s compared to 3 of 16 in the off group (p<0.05) (Koehler-McNicholas et al., 2019).

### 4.3 The Importance of Gait Speed

Gait speed is a powerful indicator of overall health and survival in the elderly population and improving gait speed is an important therapeutic goal. Based on a large population study, Studenski et al. (Studenski et al., 2011) found that gait speed, age, and gender predicted survival as well as factors related to chronic conditions, smoking history, blood pressure, and hospitalization. In fact, improvement in gait speed by 0.10 m/s was found to predict better survival in older adults (Hardy et al., 2007). Furthermore, while each decrease in gait speed by 0.10m/s has been associated with longer hospital stays and higher healthcare costs, each 0.10m/s per year increase in gait speed has been shown to be predictive of shorter hospital stays and healthcare cost reduction (Purser et al., 2005) further emphasizing the importance of gait speed as a relevant health indicator and vital sign (Middleton et al., 2015).

Healthy aging is associated with an annual decrease in gait speed by 0.013 m/s (Buracchio et al., 2010). However, it is well know that individuals with PN walk slower than their healthy counterparts (Menz et al., 2004; Lipsitz et al., 2018), likely as an adapted strategy to maintain balance (Dingwell et al., 2000). In fact, subjects with peripheral sensory loss (Lipsitz et al., 2018) who were consistently impaired over five years showed a decline in gait speed of 0.23 m/s over that time period, i.e., 0.046 m/s/yr., more than 3.5 times higher than reported by Buracchio in healthy aging (Buracchio et al., 2010). Our current population of subjects with PN, who appear similar to the “impaired” category of community-dwelling older individuals in Lipsitz et al. (Lipsitz et al., 2018), showed an increase in gait speed of 0.09 m/s following 10 weeks of use of the Walkasins device, corresponding to an annual rate of 0.47 m/s. Interestingly, the Pre-Faller group increased both their normal and fast gait speed although the effect size was smaller compared to the Pre-NonFaller group who improved only their normal gait speed.

### 4.4 Plantar Sensation and Balance Control

The improvement in gait speed and function from wearing Walkasins may be interpreted from our understanding of the sensorimotor control of human locomotion and balance. When subjects with PN perform gait and balance activities, sensory information related to foot pressure is either completely absent or at least distorted and, therefore, likely non-veridical; and it is unlikely that remaining balance-related somatosensory information can sufficiently compensate, leading to decreased stability and increased risk of balance loss. Plantar cutaneous sensory information is important for standing balance (Meyer et al., 2004b; Strzalkowski et al., 2018) gait stability (Zehr et al., 2014) signaling stance limb placement and withdrawal to facilitate phase-dependent modulation of controlling reflex responses (Zehr and Stein, 1999) as well as when responding to balance perturbations (Meyer et al., 2004a). Hlavacka et al. (Hlavacka et al., 1995) stated that an internal representation of the body vertical requires integration of somatosensory and vestibular inputs, later emphasized by Bronstein who concluded that somatosensory information has a “prominent role” in verticality perception, which is crucial for optimal balance control (Bronstein, 1999). Marsden et al. (Marsden et al., 2003) concluded that the processing of vestibular information is influenced by load-related afferent feedback for control of balance.

The integration of somatosensory and vestibular information appears of particular importance for gait function during the double support phase following foot placement during walking (Bent et al., 2004), which is near the events when Walkasins provides tactile stimuli. Although of lesser fidelity than intact plantar sensation, the Walkasins device may provide sufficient and relevant sensory information that is veridical both during standing and walking activities by signaling out of balance events during standing as well as indicating stance and swing phases of gait, which can help improve gait and balance function. Furthermore, these tactile stimuli are provided just proximally to the original sensory loss and mainly along the same dermatomes representing the plantar surface of the foot possibly making it intuitive to integrate into functional behavior (Koehler-McNicholas et al., 2019) by providing relevant sensory information to spinal central pattern generators for locomotion (Guertin, 2012).

### 4.5 Falls Data

Although we noticed an encouraging decrease in fall rate as well as in the number of fall-risk factors in the Pre-faller subgroup (Table 6), a 10-week time period with a fairly small number of subjects is too short to draw any final conclusions related to prevention of falls. Continued evaluation of longer-term data from the walk2Wellness trial during continued use of the device is currently ongoing (26 and 52 weeks) and will be reported at a future date. Interestingly, studies have found that effects of improved clinical gait and balance outcomes may lead to a delayed effect of fall reduction, which was reported following six months of Tai Chi (Li et al., 2004; Li et al., 2005) and also observed by Wolf et al. (Wolf et al., 1996; Wolf et al., 2003).

### 4.6 Participant Reported Outcomes

There appeared to be none or only minimal changes in the self-report measures throughout the 10 weeks of the trial (Table 5). Although this may seem counterintuitive, Richardson et al. (Richardson et al., 2001) reported significant improvements in clinical balance outcomes, but non-significant improvement in the ABC score following a strength and balance intervention for patients with PN. Similarly, an exercise intervention study for older adults found discrepancies between balance and ABC score improvements suggesting that the relationship between balance confidence and functional performance may not be well understood (Cyarto et al., 2008). In the current study, a lack of overall improvement in the ABC score may also be influenced by subject expectations, the short duration of study, or the time of the year as some subjects were enrolled in the winter months during snowy and icy conditions. Furthermore, results of the ABC balance confidence scores may also be viewed in the context of the subjects not being systematically informed of changes in clinical outcomes during the study, nor receiving any organized encouragement reflecting their performance.

Interestingly, upon further investigation we observed differences between the overall pattern of improvement in clinical outcomes versus self-reported measures of balance confidence as illustrated in Figure 3. Clinical outcomes at 10 weeks showed similar improvement across the full range of baseline scores, indicated by a regression line between baseline and 10-week scores being near parallel with the line of unity as illustrated with the FGA scores in Figure 3 A. However, this was not the case for the balance confidence ABC-score as seen in Figure 3 B (similar observations were made for the VADL scores, not shown here). As can be seen, the regression line between baseline and 10- week ABC scores intersects the line of unity and it has a slope of 0.47 (Figure 3 B). Interestingly, the two lines intersect at the baseline value 67%, the published cut-off value for high fall risk (Lajoie and Gallagher, 2004).

**Figure 3.**
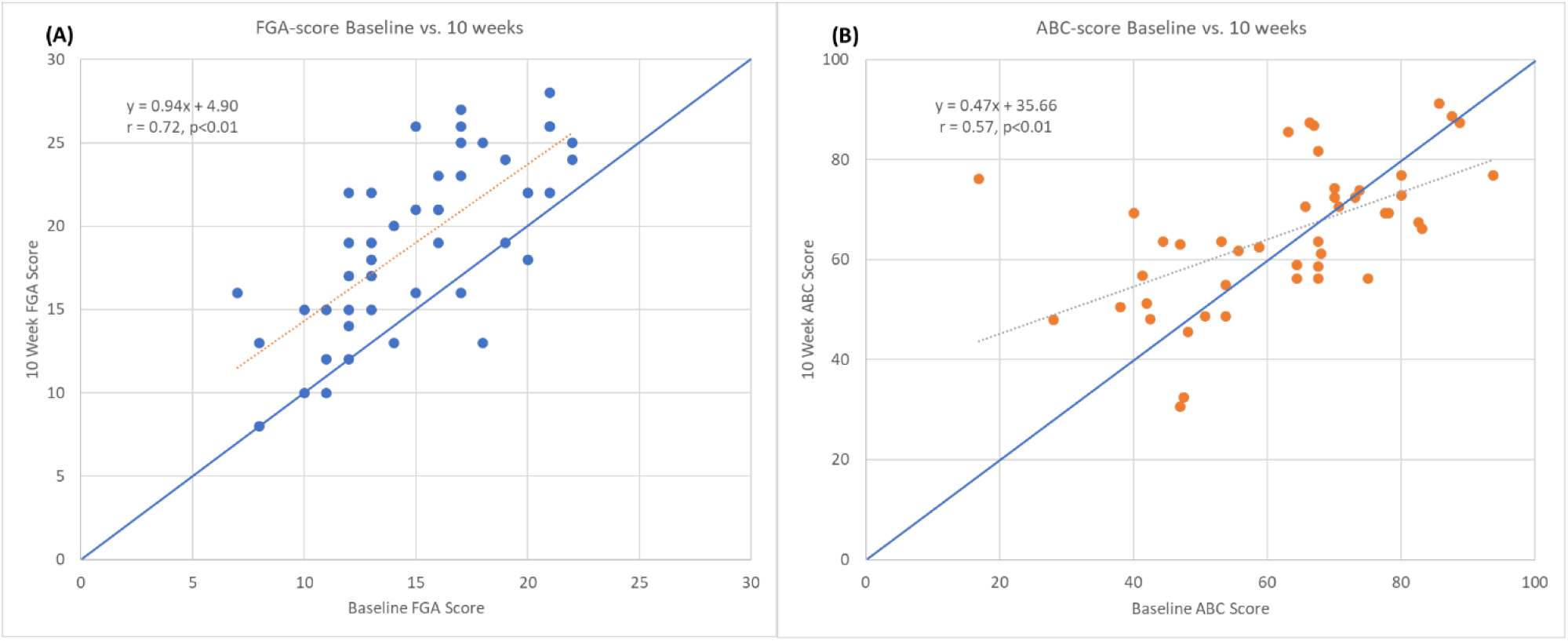
Graphs showing baseline vs. 10-week FGA (A) and ABC scores (B). Markers above line of identity indicate higher scores at 10-week assessment. Notice line of regression for FGA scores is near parallel to line of identity indicating a similar increase across all baseline FGA scores. For ABC scores the line of regression intersects the line of identify near 67% indicating an increase for lower baseline ABC scores and a decrease for higher baseline ABC scores.

Subjects enrolled in the current study were all at high fall risk based on their FGA score ≤22 (Wrisley and Kumar, 2010) as well as a diagnosis of PN (Richardson and Hurvitz, 1995). Based on additional outcomes and baseline characteristics, subjects had on average more than four fall risk factors (Table 1, Table 2, Table 6). Consequently, it could be argued that study participants with multiple impairments related to balance maybe “should not” have a balance confidence ABC score above 67% and those who report such scores are either overly confident and/or simply unaware of their true balance capabilities. Compellingly, subjects with greater balance confidence at baseline actually showed a decrease in their balance confidence during the study (from 76.5±8.1 to 71.8±9.9, p<0.02) while subjects with a baseline ABC score <67% (in the range of high-fall risk) increased their balance confidence scores (49.9±12.5% to 59.3±15.1%, p<0.01).

If we consider a simple ratio between ABC and FGA scores representing “amount” of balance confidence per FGA point, someone with 100% balance confidence and the maximum FGA score of 30 would have a ratio of 3.3. A ratio between the established cut-offs for high fall risk for ABC and FGA scores, 67% and 22, respectively, represents a ratio of 3.0. At baseline in the current trial, the overly confident subjects had a ratio of 5.1±1.3, versus 3.6±1.1 for the low confidence subjects. After 10 weeks of device use, with increased FGA scores across the board, the ratio was 3.8±1.4 for the overly confident subjects versus 3.4±1.0 for the lower confidence subjects. It appears the overly confident subjects may have “normalized” their self-perception of their balance ability while the lower confidence subjects increased their ABC score proportionally to their improved FGA score and maintained a similar self-confidence to FGA ratio. Although the FGA and ABC capture different constructs related to balance, it may be of importance for clinicians to be aware of the potential discrepancy between a patient’s self-perception and actual functional performance when developing a plan of care targeting gait, balance function and fall prevention.

The T-scores for baseline PROMIS measures (Pain Interference, Satisfaction Social Roles, Ability to Participate) were all close to 50, which was unexpected since it is considered the average for the US population (Hahn et al., 2014; Askew et al., 2016; Hahn et al., 2016a; Hahn et al., 2016b). We had expected these outcomes to deviate significantly in this complex clinical population. Consequently, any major changes in these measures should not be expected although a small statistically significant improvement in PROMIS Ability to Participate score, which remained through 10 weeks, was seen for the Pre-NonFaller sub-group from baseline to the 2-week assessment.

Some additional trends in the patient reported outcomes of interest for further research were observed, especially some differences between the Pre-Faller and Pre-NonFaller group. The Pre- Faller group had a PHQ-9 score of 5.3 at baseline, >5 considered mild depression (Kroenke et al., 2001), which decreased to 3.9 at 6 weeks and reached 4.5 at 10 weeks, changes that nearly reached overall statistical significance (p=0.08), and of minimal clinical relevance (Kroenke et al., 2001). It may be of interest to further investigate individuals with higher initial PHQ-9 scores to better understand this observation. In the Pre-NonFaller group, a non-significant (p=0.12), increasing trend in PROMIS Satisfaction Social Roles scores and a near significant improvement in VADL score (p=0.08) were seen. Finally, while VAS Pain scores were overall in the range of mild pain (≤3) and remained steady throughout 10 weeks, pain scores appeared to trend slightly lower in the Pre- NonFaller group, although not statistically significant (p=0.10) and not clinically meaningful. Here it may be of interest to further investigate individuals with higher initial pain levels, especially those with neuropathy-related foot pain by using a more disease-specific pain rating scale.

### 4.7 Study Limitations

There are several limitations to this trial. It is not blinded, lacks a control group and a placebo treatment. Unfortunately, it is not feasible to blind subjects from treatment in the current study since being able to feel the tactile stimuli from the device is an inclusion criterion. Using some form of random pattern stimuli as a sham may seem possible (Basta et al., 2011), although it is not known if such stimuli may have an effect of their own and it would not help address the question whether using the device as currently designed, according to principles of sensorimotor control of balance and gait, has an effect on gait and balance function. Consequently, the best placebo treatment would likely be wearing a device that is turned off. However, without using some form of deceit claiming the device is working although it cannot be felt, it would likely be difficult to recruit participants for such research and/or to ensure long-term compliance. In addition, incorporating a minimal stimulation amplitude as a sham, assuming it has no effect may be incorrect since studies implementing stochastic resonance using subsensory mechanical noise have demonstrated improvement in balance (Lipsitz et al., 2015). Furthermore, using a randomized control cross-over design, we recently demonstrated in-clinic improvements in clinical outcomes when the Walkasins device was worn and turned on as compared to turned off (Koehler-McNicholas et al., 2019).

Consequently, we felt comfortable incorporating a single treatment arm design knowing the in-clinic effects. Furthermore, any placebo effects were likely decreased by not systematically informing subjects about any changes in outcomes and minimizing encouragement during interactions with subjects that could affect expectation and beliefs in the treatment (Finniss et al., 2010; Enck et al., 2013; Coste and Montel, 2017), and prohibiting any additional balance training/therapy intervention during the 10 weeks of the trial.

If the effects in this study were placebo our findings should align with research findings on the placebo arm of randomized, placebo-controlled trials (Wartolowska et al., 2016). A systematic review of temporal changes in the placebo arm across 47 surgical randomized control trials found that effects size of subjective outcomes was large (0.64), while effect size of objective clinical outcomes was small (0.11) (Wartolowska et al., 2016). Furthermore, major differences in placebo-effect sizes have been reported with subject-reported self-perception effects being larger than observer-based ratings (Rief et al., 2009). On the contrary, effect sizes in the current study were large for the clinical outcomes and small for the self-reported outcomes, supporting the interpretation that effects were due to device use and not placebo. Further support of this view includes relatively high subject compliance and reported device use and, a low subject dropout rate as well as the sustained duration and continued gradual improvement in clinical outcomes throughout the 10-week period. An additional weakness includes enrollment of mostly male subjects, partly due to nearly half of the subjects being Veterans, who especially in this older generation are predominantly male. Strengths of this trial include involvement of multiple sites across different geographies with different assessors at different clinics limiting confounding balance interventions, and the use of standardized objective clinical outcome measures.

## 5 Conclusion

A wearable sensory neuroprosthesis may provide a new way to treat gait and balance problems and manage falls in high fall-risk patients with PN. Longer term data would be required to further investigate actual decreases in falls.

## 6 Acknowledgments

We thank Lori Danzl, Alexandria Lloyd, Christine Olney (Minneapolis Department of Veterans Affairs Health Care System, Minneapolis, MN), Tien Dat Nguyen, (M Health Fairview, Minneapolis, MN), Nathan Silver (Baylor College of Medicine, Houston, TX), Ikechukwu Iloputaife, Wanting Yu (Hinda and Arthur Marcus Institute for Aging Research, Hebrew SeniorLife, Roslindale, MA) for providing study coordination, subject recruitment and data gathering activities. We thank Annette Xenopoulos-Oddsson (University of Minnesota, MN,), Dr. Lee Newcomer and Dr. Steven Stern for reviewing drafts of the manuscript. The manuscript was published as a MedRxiv preprint (Oddsson et al., 2020b).

## 7 Contribution to the Field Statement

Peripheral neuropathy is a condition affecting over 20 Million individuals in the US alone that causes damage to nerves in the peripheral nervous system. Peripheral neuropathy is often related to diabetes, chemotherapy treatment or has unknown causes. Symptoms of the disease that involve numbness and loss of sensation in the feet often lead to problems with gait and balance and a high risk of falls. We report results from the multi-site clinical trial, walk2Wellness, that has investigated long-term effects of daily use of a new wearable sensory neuroprosthesis on gait function, balance, fall rates, and quality of life in a group of peripheral neuropathy patients. The device (Walkasins®, RxFunction Inc., MN, USA) provides directional tactile stimuli around the ankle during standing and walking activities reflecting changes in foot pressure related to balance. Patients can then “feel” their feet in contact with the ground, notice body sway during standing replacing their lost afferent nerve function. Our results after 10 weeks of daily use show improved walking balance, speed of walking, ability to rise from a chair move around and sit down and an encouraging trend in lower fall rates. Also, participants with initial low self-confidence in their balance showed higher self-confidence.

## Notes

### Competing Interest Statement

Mixed competing interests: All authors have completed the ICMJE uniform disclosure form at www.icmje.org/coi_disclosure.pdf and declare: LO is an inventor of the technology, Co-Founder of RxFunction, a shareholder in the company, serves on its Board of Directors and is an employee of the company. LJ is an employee of RxFunction and YR provides paid consulting services to RxFunction. The remaining authors received grant funding from RxFunction for the submitted work; no financial relationships with any organizations that might have an interest in the submitted work in the previous three years; no other relationships or activities that could appear to have influenced the submitted work.

### Clinical Trial

NCT03538756

### Clinical Protocols

https://clinicaltrials.gov/ct2/show/NCT03538756

### Funding Statement

The walk2Wellness trial is completely funded by RxFunction. Sites have received funding from RxFunction to conduct research activities related to the trial.

### Author Declarations

Human subject testing was approved according to the Declaration of Helsinki by Advarra IRB (formerly Quorum Review IRB), serving as the Institutional Review Board (IRB) of record for three of the participating sites under the study protocol. The three sites include Baylor College of Medicine, Houston, TX; Hebrew SeniorLife, a Harvard Medical School Affiliate, Boston, MA; and M Health Fairview, Minneapolis, MN. The IRB Subcommittee, the Subcommittee on Research Safety, and the Research and Development Committee of the Minneapolis VA Health Care System (MVAHCS) also approved the trial. The study is registered on ClinicalTrials.gov (#NCT03538756).

### Summary of Updates

Corrected author order. Revised Acknowledgement. Minor clarifications.

